# Proactive social distancing mitigates COVID-19 outbreaks within a month across 58 mainland China cities

**DOI:** 10.1101/2020.04.22.20075762

**Authors:** Zhanwei Du, Xiaoke Xu, Lin Wang, Spencer J. Fox, Benjamin J. Cowling, Alison P. Galvani, Lauren Ancel Meyers

**Affiliations:** The University of Texas at Austin, Austin, Texas 78712, The United States of America; Dalian Minzu University, Dalian 116600, China; University of Cambridge, Cambridge CB2 3EH, UK; The University of Hong Kong, 7 Sassoon Rd, Hong Kong SAR, China; Center for Infectious Disease Modeling and Analysis, Yale School of Public Health, New Haven, Connecticut, USA; Santa Fe Institute, Santa Fe, New Mexico, The United States of America

**Author notes:** Correspondence: Lauren Ancel Meyers.

**Keywords:** coronavirus, epidemiology, reproduction number, non-pharmaceutical interventions

## Abstract

In early 2020, cities across China enacted strict social distancing measures to contain emerging coronavirus (COVID-19) outbreaks. We estimated the speed with which these measures contained community transmission in each of 58 Chinese cities. On average, containment was achieved 7.83 days (SD 6.79 days) after the implementation of social distancing interventions, with an average reduction in the reproduction number (*R*_*t*_) of 54.3% (SD 17.6%) over that time period. A single day delay in the implementation of social distancing led to a 2.41 (95% CI: 0.97, 3.86) day delay in containment. Swift social distancing interventions may thus achieve rapid containment of newly emerging COVID-19 outbreaks.

---

On December 31, 2019, a cluster of atypical pneumonia in Wuhan, China was reported to the regional office of the World Health Organization (WHO). Its etiology was later identified as the novel SARS-CoV-2 coronavirus. The disease (COVID-19) spread rapidly across China and internationally ^1^. As of April 9, 2020, there have been 1,436,198 confirmed cases and 85,522 deaths in 209 countries ^2^. In the absence of pharmaceutical prophylactic options, the primary means of COVID-19 control are social distancing interventions, including school closures, work restrictions, shelter-in-place measures and travel bans. In late January, reported cases rose steeply in the Hubei province of China and imported cases sparked outbreaks in many other cities throughout China. By February 14, 2020, the government had limited the movement of over 500 million people across 80 cities ^3,4^. During this time, many of these cities rapidly enacted multiple social distancing orders to slow the local spread of the virus, such as restricting non-essential services and public transit ^5–8^. Given the substantial economic and societal costs of such measures ^9^, estimates of their effectiveness can serve as critical evidence for intervention policy decisions worldwide ^10,11^.

Using case data from online reports published by the China CDC and health commissions, we estimated the time elapsed between the first reported case in a city and successful containment of the outbreak (χ). Technically, we consider an outbreak contained when the 95% confidence interval of the instantaneous reproduction number (*R*_t_) drops below one. We analyzed the speed of COVID-19 containment for 58 cities in Mainland China outside of Hebei Province with at least 20 confirmed cases by February 14, 2020 (**Figure 1** and **Appendix Table S2**). Collectively, these cities deployed seven different types of interventions over the course of their epidemics ^12^: (1) bans on entertainment and public gatherings; (2) broad restrictions on public service including healthcare, schooling, shopping and restaurants; (3) initiation of a level-1 response entailing systematic testing and isolation of confirmed cases; (4) suspension of intra-city public transport; (5) suspension of travel between cities; (6) reporting of confirmed cases; (7) recruitment of governmental staff and volunteers to enforce quarantine and social distancing. On average, the first social distancing measure was enacted 13 days (95% CI: 3, 22.05) following the first confirmed case. At the time of the first measure, the 58 cities had an average of 40 (95% CI: 9, 209) cumulative reported cases. The cities achieved containment an average of 21 days (95% CI: 7.95, 35.1) after the first reported case and 11.5 days (95% CI: −5.1, 25.2) following implementation of the earliest interventions. During the period of containment, the reproduction number (*R*_t_) declined by an average of 54.3% (SD 17.6%) (**Appendix Figure S2**).

**Figure 1.**
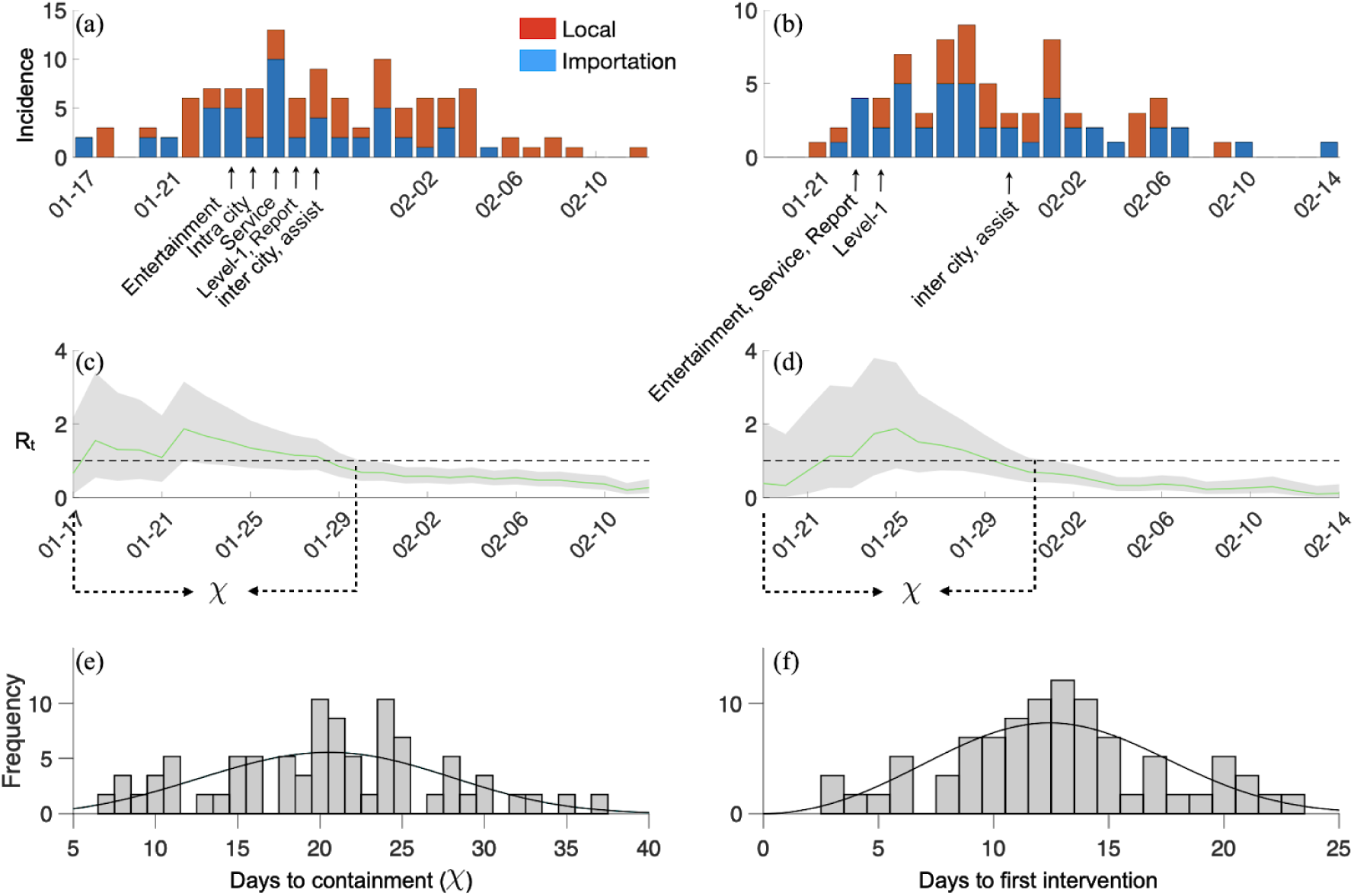
COVID-19 introductions, transmission and containment in Mainland China cities prior to February 15, 2020. We demonstrate the methodology for two provincial capitals: (a, c) Xi’an and (b, d) Nanjing. (a,b) The estimated daily incidence of imported and locally-transmitted COVID-19 cases and the implementation times for local social distancing measures. (c,d) The estimated daily time-varying reproduction numbers (*R*_t_), with the green line and shading indicating the median and 95% credibility bounds for *R*_t_, respectively. We calculate the number of days from the first reported imported case until the upper 95% CI of *R*_t_ drops below one (χ) for 58 cities in Mainland China. (e) The distribution of χ across 58 cities in Mainland China (also see Appendix Figure S2). The average duration of outbreaks is 21 days (95% CI: 7.95, 35.1). Based on an AIC comparison between Gamma, Lognormal and Weibull distributions fitted via maximum likelihood to the data (Appendix Table S3), we find that the χ are roughly Weibull distributed with scale 22.94 (95% CI: 21.12, 24.91) and shape 3.28 (95% CI: 2.68, 4.02), as indicated by black line. (f) The distribution of time between the first locally reported case and the first social distancing measure resembles a Weibull distribution with scale 14.24 (95% CI: 13.01,15.60) and shape 2.98 (95% CI: 2.44,3.65).

Using a combination of linear regression and best-subsets model selection ^13^, we found that the timing of the first intervention and the initiation of level-1 response significantly predicted the speed of containment across the 36 cities that deployed all seven interventions (*R*^2^=0.27, *p* < 0.001) (**Appendix Figure S1**). A delay of a single day in implementing the first intervention is expected to prolong an outbreak by 2.41 (95% CI: 0.96, 3.86) days. In contrast, the timing of the level-1 response was inversely related to the speed of containment. Level-1 responses were initiated by the central government across Mainland China over the course of one week, starting with the hardest hit areas in and near Hubei Province on the first day and working outwards towards more distant cities. Thus, the day of Level-1 initiation within this one-week period is likely an indicator for the initial severity of an outbreak and the corresponding difficulty of containment.

We have estimated the value of *proactive* social distancing interventions in terms of a reduction in days until successful containment. However, since most cities implemented multiple measures quickly and simultaneously, we are unable to disentangle the efficacies of individual modes of social distancing. We also note that our estimates of *R*_t_ may be biased by the limited case report data available prior to February 14, 2020; we lack information about testing rates and priorities in China during this period. As public health agencies around the globe struggle to determine when to implement potentially costly social distancing measures, these estimates highlight the potential long-term benefits of early and decisive action.

## Data Availability

Not applicable

## Acknowledgements

We thank Dr. Simon Cauchemez for helpful discussions. We acknowledge the financial support from NIH (U01 GM087719) and the National Natural Science Foundation of China (61773091).

